# Factors associated with circulating levels of IL-17A in a high HIV-burden population

**DOI:** 10.64898/2025.12.16.25342346

**Authors:** David Chisompola, Emmanuel Luwaya, Joseph M. Chalwe, Joreen P. Povia, Annet Kirabo, Sepiso K. Masenga

**Affiliations:** HAND Research Group, School of Medicine and Health Sciences, Mulungushi University, Livingstone 10101, Zambia; Department of Cardiovascular Science and Metabolic Diseases, Livingstone Center for Prevention and Translational Science, Livingstone 10101, Zambia; Nuclear Medicine Research Infrastructure (NuMeRI), Steve Biko & Malan Street, Capital Park, Pretoria, 0084, South Africa; Department of Nuclear Medicine, University of Pretoria & Steve Biko Academic Hospital, Pretoria 0001, South Africa; Department of Health Economics, Livingstone Center for Prevention and Translational Science, Livingstone 10101, Zambia; Department of Medicine, Vanderbilt University Medical Center, Nashville, TN 37203, USA; Vanderbilt Center for Immunobiology, Vanderbilt Institute for Infection, Immunology and Inflammation, Vanderbilt University Medical Center, Nashville, TN 37203, USA; Department of Molecular Physiology and Biophysics, Vanderbilt University Medical Center, Nashville, TN 37203, USA; Vanderbilt Institute for Global Health, Vanderbilt University Medical Center, Nashville, TN 37203, USA

**Keywords:** Interleukin-17A, HIV, Inflammation, Potassium, Metabolic Syndrome, Cross-Sectional Studies

## Abstract

**Background:** Interleukin-17A (IL-17A) is a key cytokine in inflammation and autoimmunity. However, its systemic correlates, particularly in a high HIV burden population, are not fully elucidated. This study aimed to identify the sociodemographic, clinical, inflammatory, metabolic, and renal factors independently associated with plasma IL-17A levels.

**Methods:** This cross-sectional analysis enrolled a cohort of adults from Livingstone Teaching Hospital in Zambia. Sociodemographic and clinical variables were systematically collected. Associations between plasma IL-17A levels and a range of covariates were assessed using simple and multiple linear regression analyses in StatCrunch to identify independent correlates. Statistical significance was set at p<0.05.

**Results:** A cohort of 366 participants was analyzed, characterized by a predominance of females (70.2%) and people living with HIV (73.5%), the vast majority of whom were receiving integrase strand transfer inhibitors (INSTI)-based antiretroviral therapy. Initial bivariate analysis revealed weak associations between IL-17A and various metabolic and inflammatory markers. In the final multivariable model (Model 1), which included all significant univariate variables, five factors were independently associated with IL-17A: IFN-γ (β: 1.17, 95% CI: 1.01–1.33, p<0.0001), IL-1 (β: 1.60, 95% CI: 0.74–2.47, p=0.0004), IL-5 (β: −0.34, 95% CI: −0.65 – −0.040, p=0.0268), soluble ST2 (β: −26.01, 95% CI: −38.15 – −13.87, p<0.0001), and D-dimer (β: −0.006, 95% CI: −0.009 – −0.002, p=0.0012). Plasma potassium was not significant in this full model (β: 0.78, 95% CI: −3.99–5.56, p=0.744). HIV status was not an independent correlate (β: 95.73, 95% CI: −499.68–691.15, p=0.749). A second model (Model 2) was constructed by adjusting for HIV status only. In this model, plasma potassium was a significant independent correlate of IL-17A (β: 10.07, 95% CI: 4.39–15.76, p=0.0006), along with triglycerides, LDL cholesterol, VLDL, IL-6, TNF-α, IL-1, IL-5, soluble ST2, fasting glucose, and lymphocyte count.

**Conclusion:** In a comprehensive model, inflammatory markers (IFN-γ, IL-1, IL-5, sST2) and D-dimer were the independent correlates of IL-17A. The relationship between plasma potassium and IL-17A was context-dependent, significant only in a model excluding other inflammatory cytokines. HIV status was not independently associated with IL-17A when inflammatory markers were considered. These findings highlight complex immune interactions and warrant further investigation.

## Introduction

Interleukin-17A (IL-17A) is a pro-inflammatory cytokine produced primarily by T helper 17 (Th17) cells and is recognized as a mediator linking immune dysregulation to a broad spectrum of chronic diseases [1]. Its production is potently stimulated by upstream cytokines, including IL-1β and IL-6, which promote the differentiation and expansion of Th17 cells [2]. While the role of IL-17A in conditions like psoriasis and rheumatoid arthritis is well-defined, its regulation in the context of chronic viral infections and associated metabolic disturbances is less understood.

Despite effective antiretroviral therapy (ART), Human Immunodeficiency virus (HIV) infection persists as a state of chronic immune activation and CD4+ T-cell dysregulation [3]. A key feature of this dysregulation is the disruption of the Th17/regulatory T cell (Treg) balance, which compromises mucosal integrity and fuels systemic inflammation [4]. This persistent inflammatory state is a critical driver of non-AIDS-defining comorbidities, including metabolic and renal diseases [5]. With the global shift towards integrase strand transfer inhibitors (INSTI)-based ART regimens, which have distinct immunomodulatory effects, there is a need to re-evaluate drivers of inflammation, such as IL-17A, in contemporary treated HIV infection.

Epidemiological evidence supports that people living with HIV have a higher prevalence and faster progression of chronic kidney disease (CKD) compared with uninfected individuals, driven by the interplay of chronic inflammation, metabolic dysfunction, and direct viral and treatment-related kidney injury [5–8]. This synergy creates a significant public health challenge by increasing morbidity and mortality in this population

Beyond infectious disease, emerging evidence suggests IL-17A is intertwined with metabolic homeostasis. It has been linked to insulin resistance, atherosclerosis, and, intriguingly, kidney function, where it can promote fibrosis and inflammation [9,10]. However, most studies investigate IL-17A in isolation. A comprehensive analysis examining its simultaneous relationship with sociodemographic, clinical, inflammatory, and metabolic factors, particularly in a population with a high burden of chronic inflammation like people living with HIV, is lacking.

Therefore, this study aimed to identify the sociodemographic, clinical, inflammatory, metabolic, and renal factors independently associated with plasma IL-17A levels in a high HIV-burden population. We hypothesized that IL-17A levels would be associated not only with HIV status and specific inflammatory cytokines but also with distinct metabolic parameters, potentially unveiling novel interactions between immunity and systemic physiology.

## Materials and Methods

### Study Design and Population

This was a cross-sectional study involving 366 adults recruited from the outpatient medical clinical at Livingstone University Teaching Hospital who were attending routine medical checkup between 01/10/2023 and 01/06/2024.

### Eligibility Criteria

Eligible participants were adults aged 18 years or older, provided written informed consent before enrolling on the study, living with (PWH) or without HIV (PWTH) and hypertension. Those with HIV were on stable antiretroviral therapy for at least 12 months and were virologically suppressed (viral load <50 copies/mL). People without HIV had a negative HIV test. To minimize confounding effects on inflammatory markers, we excluded individuals with diabetes, pregnancy, severe renal or hepatic disease, active opportunistic infections, or other serious comorbidities.

### Sample Size and Sampling technique

A purposive sampling technique was employed to recruit participants. This non-probability method was chosen to efficiently enroll as many participants as possible. A sample size calculation was performed for a multiple linear regression model. To detect a medium effect size (f^2^ = 0.15) with a significance level (α) of 0.05 and a statistical power of 90% for a model testing up to 12 predictors, a minimum sample size of n = 158 was required [11]. To account for potential data issues and to ensure robust subgroup analysis, we targeted a larger sample. The final recruited cohort of n=366 participants therefore provided ample statistical power to reliably identify the independent correlates of IL-17A reported in this analysis.

### Sociodemographic and Clinical Data

Sociodemographic and clinical data were systematically collected and managed using Research Electronic Data Capture (REDCap) platform. Trained research personnel gathered information on age, sex, marital status, employment status, and medical history (including HIV, hypertension, and cardiovascular conditions) through standardized structured interviews and medical record reviews. HIV status and ART regimen details were confirmed from clinical records.

### Biochemical Measurements

Following an overnight fast, venous blood samples were collected. Plasma was separated and stored at −80°C until analysis. IL-17A, IL-1, IL-5, IL-6, TNF-α, d-dimer and IFN-γ were quantified using an enzyme-linked immunosorbent assay (ELISA) commercially available kits, following manufacturer protocols. Fasting glucose, lipid profile (triglycerides, LDL-cholesterol, HDL-cholesterol, VLDL), and plasma potassium were measured using standard automated clinical chemistry analyzers.

### Statistical Analysis

Data were exported from the REDCap platform into a structured format and underwent initial cleaning in Microsoft Excel to ensure data integrity. All statistical analyses were performed using StatCrunch. Descriptive statistics were presented as medians with interquartile ranges (IQR) for continuous variables and frequencies with percentages for categorical variables. The relationship between IL-17A and covariates was first assessed using simple linear regression, with results visualized in a correlogram and scatter plots. Variables showing a significance level of p < 0.05 in the univariate analysis were included in the multiple linear regression models. Two models were constructed:

**Model 1:** Included all significant variables from univariate analysis.

**Model 2:** A restricted model was built by adjusting for HIV status alone to assess the independent association of other covariates in a clinically oriented context.

A p-value of < 0.05 was considered statistically significant in the multivariable models.

### Ethical considerations

Ethical approval for the study was obtained from the Mulungushi University School of Medicine and Health Sciences Research Ethics Committee on 09 July 2023 (Ref. SMHS-MU3-2023-005), with additional administrative clearance granted by Livingstone University Teaching Hospital. Written informed consent was obtained from all participants prior to enrollment. The study was conducted in accordance with the principles of the Declaration of Helsinki and relevant national ethical guidelines. All data were anonymized at the point of collection, and no information capable of identifying individual participants was recorded or retained.

## Results

### Sociodemographic and Clinical Characteristics of the Cohort

The cohort was predominantly female (70.2%), with a median age of 48.5 (40.0–59.0, IQR) years, and had a high prevalence of HIV (73.5%) (Table 1). Most participants were on an INSTI-based ART regimen (95.8%) and reported good ART adherence (71.9%). While common comorbidities included hypertension (24.3%) (Table 1).

**Table 1.**
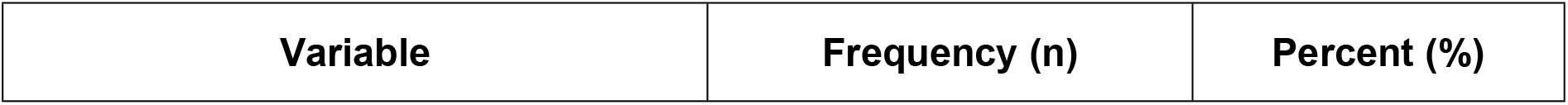
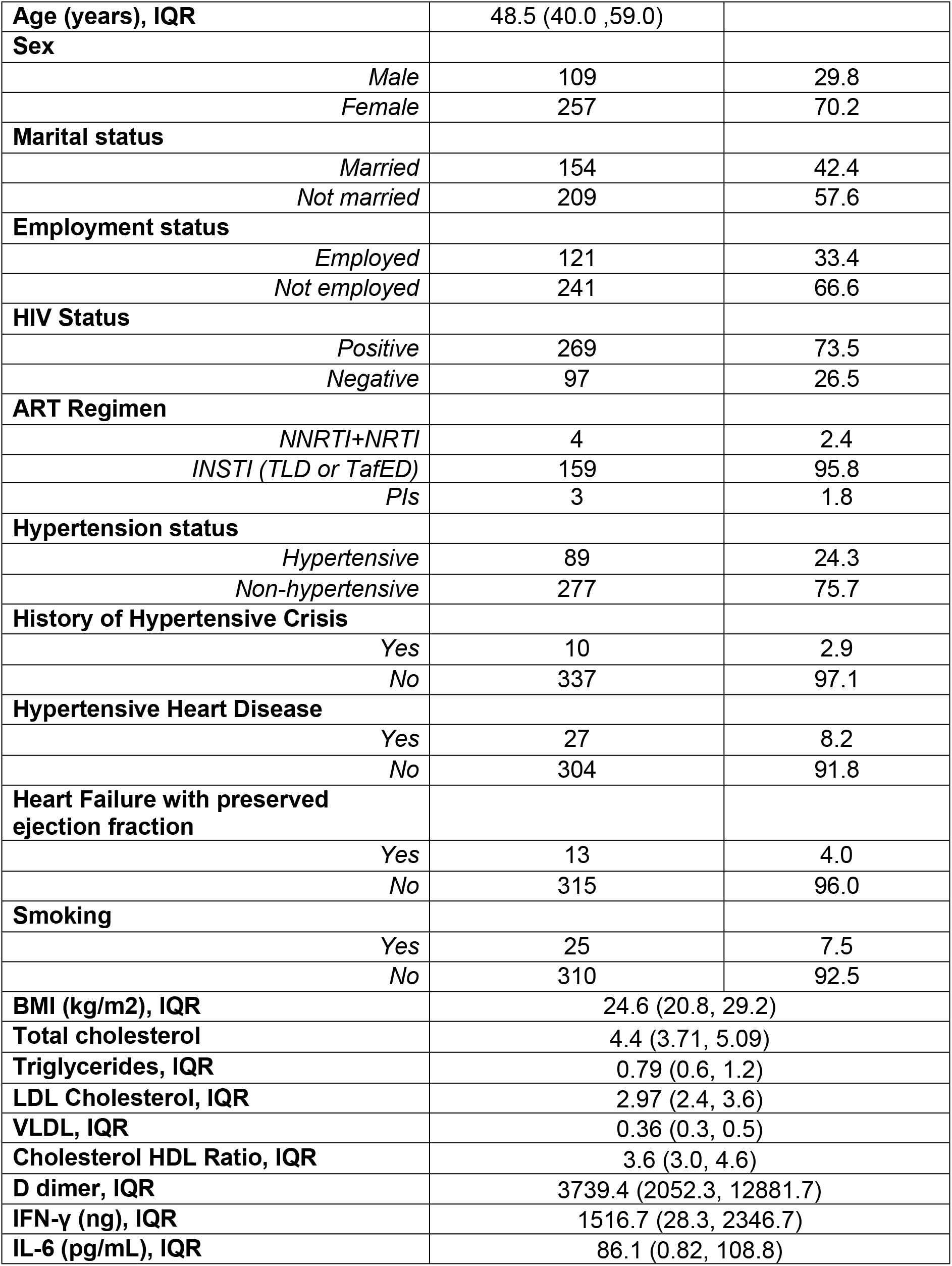
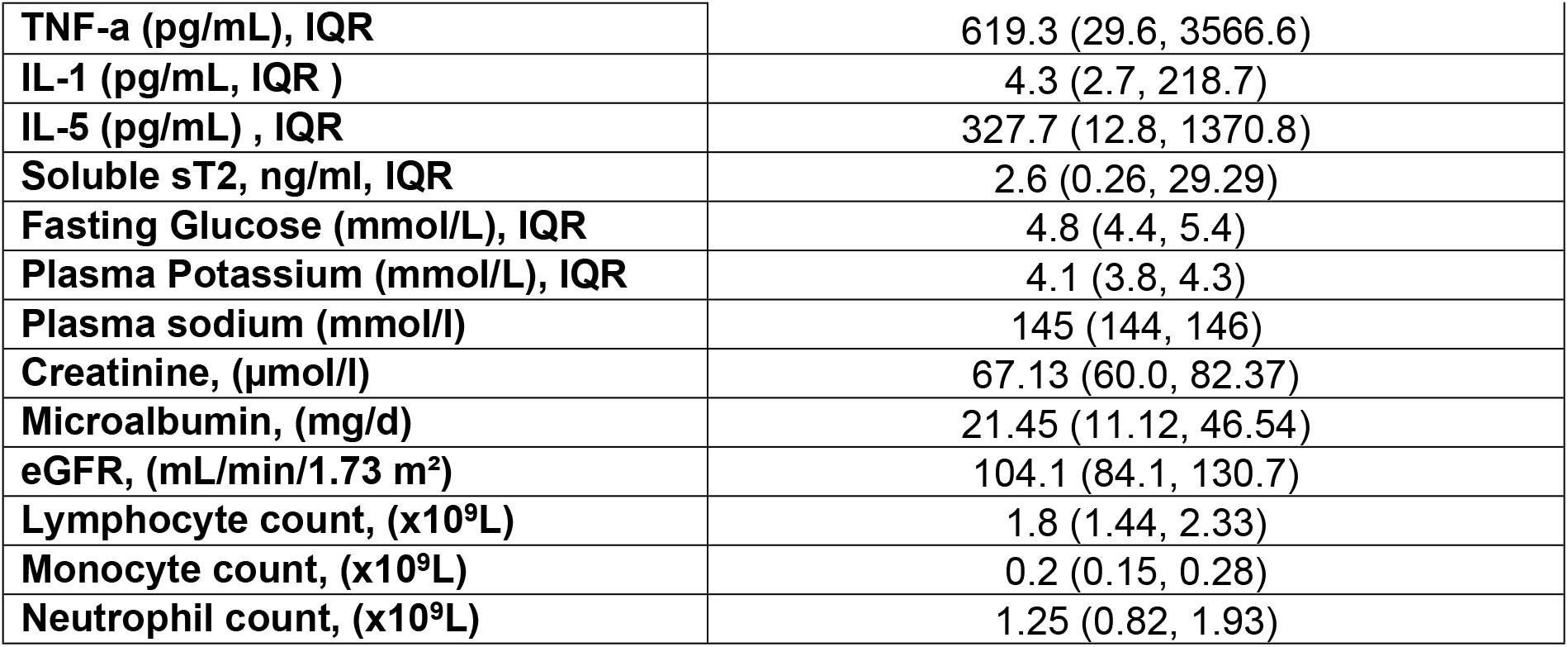
Participants Demographic Sociodemographic, clinical, inflammatory and metabolic and kidney correlates of IL-17a.

| Variable | Frequency (n) | Percent (%) |
| --- | --- | --- |
| **Age (years), IQR** | 48.5 (40.0 ,59.0) |  |
| **Sex** |  |  |
| *Male* | 109 | 29.8 |
| *Female* | 257 | 70.2 |
| **Marital status** |  |  |
| *Married* | 154 | 42.4 |
| *Not married* | 209 | 57.6 |
| **Employment status** |  |  |
| *Employed* | 121 | 33.4 |
| *Not employed* | 241 | 66.6 |
| **HIV Status** |  |  |
| *Positive* | 269 | 73.5 |
| *Negative* | 97 | 26.5 |
| **ART Regimen** |  |  |
| *NNRTI+NRTI* | 4 | 2.4 |
| *INSTI (TLD or TafED)* | 159 | 95.8 |
| *PIs* | 3 | 1.8 |
| **Hypertension status** |  |  |
| *Hypertensive* | 89 | 24.3 |
| *Non-hypertensive* | 277 | 75.7 |
| **History of Hypertensive Crisis** |  |  |
| *Yes* | 10 | 2.9 |
| *No* | 337 | 97.1 |
| **Hypertensive Heart Disease** |  |  |
| *Yes* | 27 | 8.2 |
| *No* | 304 | 91.8 |
| **Heart Failure with preserved ejection fraction** |  |  |
| *Yes* | 13 | 4.0 |
| *No* | 315 | 96.0 |
| **Smoking** |  |  |
| *Yes* | 25 | 7.5 |
| *No* | 310 | 92.5 |
| **BMI (kg/m2), IQR** | 24.6 (20.8, 29.2) |  |
| **Total cholesterol** | 4.4 (3.71, 5.09) |  |
| **Triglycerides, IQR** | 0.79 (0.6, 1.2) |  |
| **LDL Cholesterol, IQR** | 2.97 (2.4, 3.6) |  |
| **VLDL, IQR** | 0.36 (0.3, 0.5) |  |
| **Cholesterol HDL Ratio, IQR** | 3.6 (3.0, 4.6) |  |
| **D dimer, IQR** | 3739.4 (2052.3, 12881.7) |  |
| **IFN-γ (ng), IQR** | 1516.7 (28.3, 2346.7) |  |
| **IL-6 (pg/mL), IQR** | 86.1 (0.82, 108.8) |  |

|  |  |
| --- | --- |
| TNF-a (pg/mL), IQR | 619.3 (29.6, 3566.6) |
| IL-1 (pg/mL, IQR ) | 4.3 (2.7, 218.7) |
| IL-5 (pg/mL) , IQR | 327.7 (12.8, 1370.8) |
| Soluble sT2, ng/ml, IQR | 2.6 (0.26, 29.29) |
| Fasting Glucose (mmol/L), IQR | 4.8 (4.4, 5.4) |
| Plasma Potassium (mmol/L), IQR | 4.1 (3.8, 4.3) |
| Plasma sodium (mmol/l) | 145 (144, 146) |
| Creatinine, (μmol/l) | 67.13 (60.0, 82.37) |
| Microalbumin, (mg/d) | 21.45 (11.12, 46.54) |
| eGFR, (mL/min/1.73 m<sup>2</sup>) | 104.1 (84.1, 130.7) |
| Lymphocyte count, (x10<sup>9</sup>L) | 1.8 (1.44, 2.33) |
| Monocyte count, (x10<sup>9</sup>L) | 0.2 (0.15, 0.28) |
| Neutrophil count, (x10<sup>9</sup>L) | 1.25 (0.82, 1.93) |

### Correlogram of IL-17A covariates

Among the continuous variables, IL-17A only correlated positively and significantly with IFN-γ, d-dimer, IL-6, TNF-a, IL-1, IL-5, soluble sT2, lymphocyte count, plasma potassium, microalbumin, triglycerides, LDL cholesterol, VLDL, LDL:HDL ratio, and total cholesterol: HDL ratio, p<0.05, **Figure 1**.

**Figure 1.**
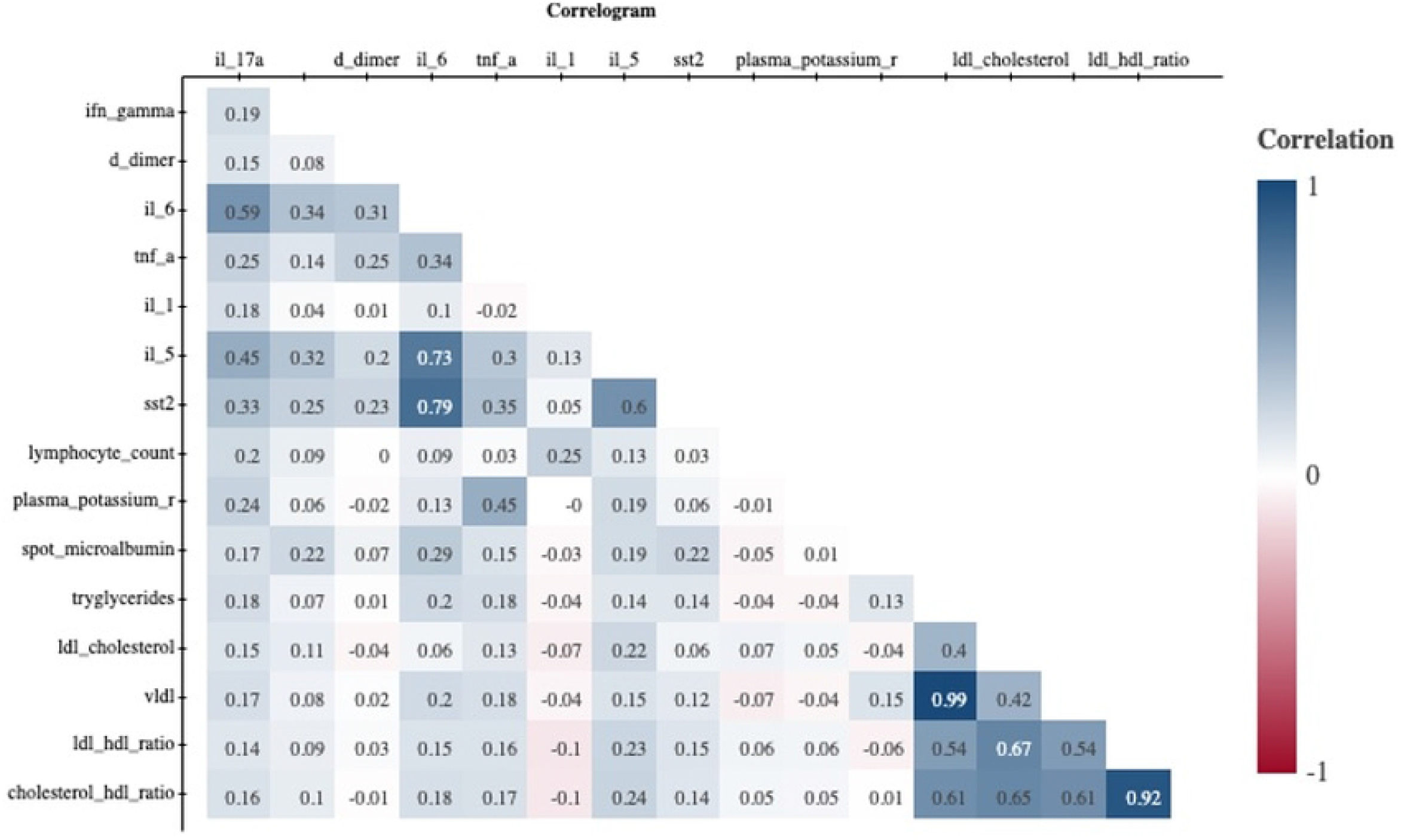
Correlogram of IL-17A covariates. IFN-γ, interferon-gamma; D-dimer, fibrin degradation product (D-dimer); TNF-α, tumor necrosis factor-alpha; Soluble sST2, soluble suppression of tumorigenicity-2; LDL cholesterol, low-density lipoprotein cholesterol; VLDL, very-low-density lipoprotein; LDL:HDL, low-density lipoprotein to high-density lipoprotein ratio

### Relationship between Il-17A and covariates in simple linear regression

Simple linear regression analyses revealed statistically significant positive correlations between plasma IL-17A levels and a range of clinical, inflammatory, metabolic, and renal variables (Fig. 2 A–L and Fig. 3 A-L). Although patterns varied across parameters, most variables demonstrated weak or inconsistent relationships with IL-17A, indicating considerable inter-individual variability. IL-17A was positively associated with living with HIV (Fig 2A), triglycerides (Fig 2E), LDL cholesterol (Fig 2F), VLVL (Fig 2G), cholesterol-to-HDL ratio (Fig 2H), d-dimer (Fig 2I), IFN-gamma (Fig 2J), IL-6 (Fig 2K), TNF-alpha (Fig 2L), IL-1 (Fig 3A), IL-5 (Fig 3B), soluble ST2 (Fig 3C), fasting blood glucose (Fig 3D), plasma potassium (Fig 3E), microalbumin (Fig 3H), and lymphocyte (Fig 3J).

**Fig 2.**
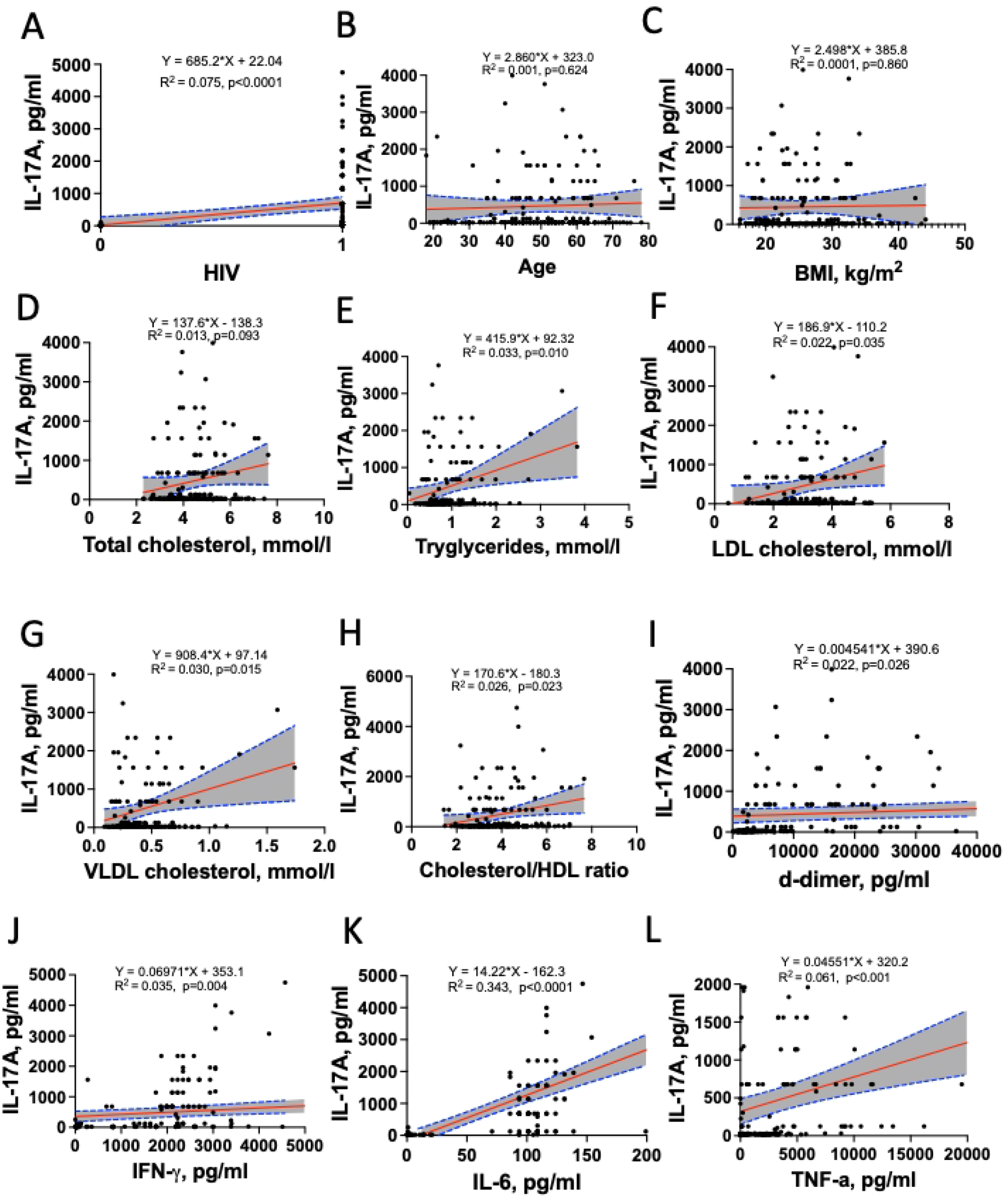
Relationship between Il-17A and lipids, demographic, anthropometric and inflammation markers in simple linear regression.

**Fig 3.**
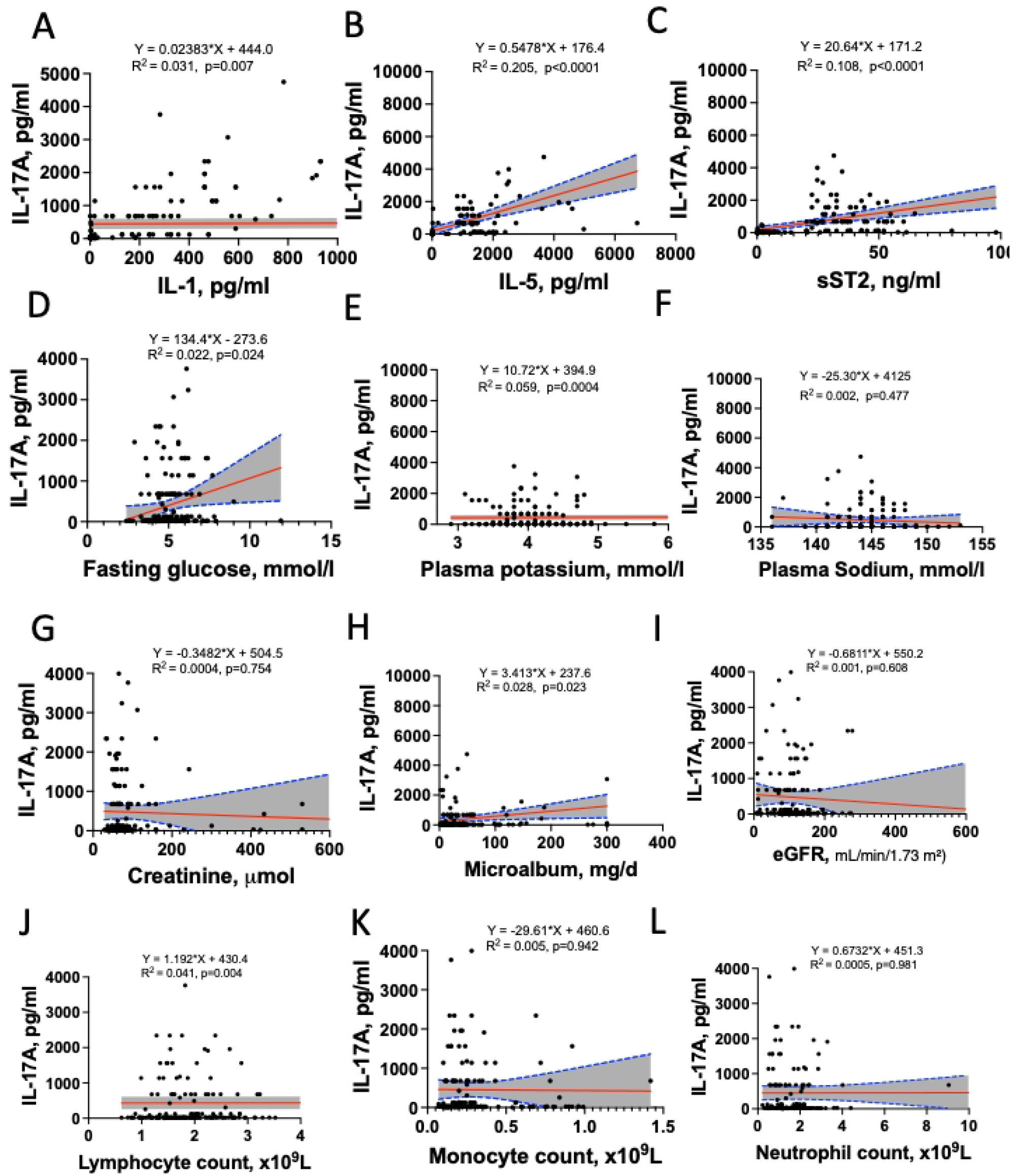
Relationship between Il-17A and inflammation, immune cells, kidney and metabolic markers in simple linear regression.

### Multiple Linear Regression Analysis of Factors Associated with IL-17A

To assess the independent associations of clinical, metabolic, and inflammatory factors with IL-17A, we constructed two multivariable regression models.

In Model 1, which included all candidate variables significant in univariate analysis, five factors were independently associated with IL-17A (Table 2). Positive correlates were IFN-γ (β: 1.17, 95% CI: 1.01–1.33, p<0.0001) and IL-1 (β: 1.60, 95% CI: 0.74–2.47, p=0.0004). Negative correlates were IL-5 (β: −0.34, 95% CI: −0.65 – −0.040, p=0.0268), soluble ST2 (β: −26.01, 95% CI: −38.15 – −13.87, p<0.0001), and D-dimer (β: −0.006, 95% CI: −0.009 – −0.002, p=0.0012). Plasma potassium (β: 0.78, 95% CI: −3.99–5.56, p=0.744) and HIV status (β: 95.73, 95% CI: −499.68–691.15, p=0.749) were not statistically significant in this model.

**Table 2.**
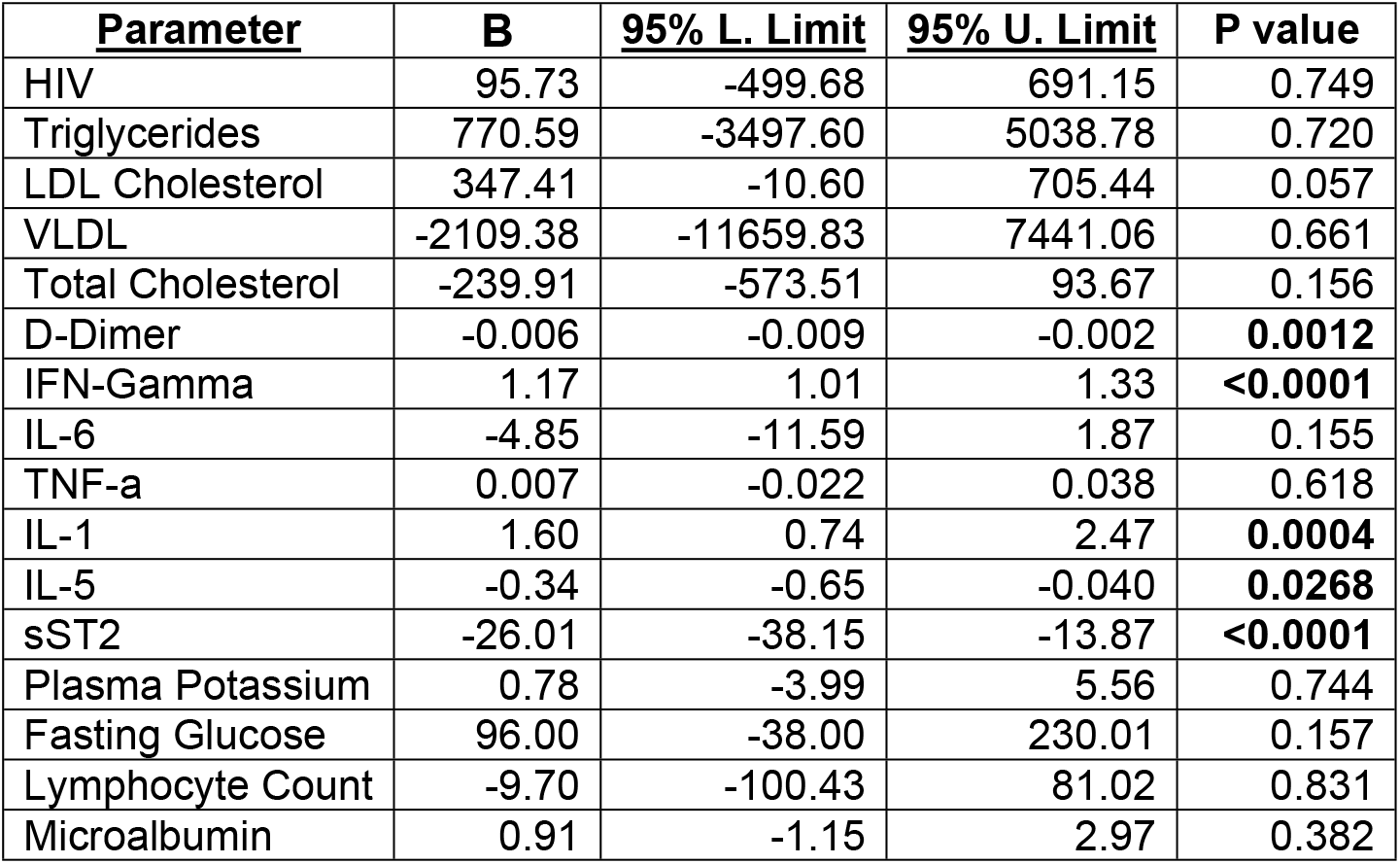
Multiple linear analysis of factors associated with IL-17A model 1.

| **Parameter** | **B** | **95% L. Limit** | **95% U. Limit** | **P value** |
| --- | --- | --- | --- | --- |
| HIV | 95.73 | -499.68 | 691.15 | 0.749 |
| Triglycerides | 770.59 | -3497.60 | 5038.78 | 0.720 |
| LDL Cholesterol | 347.41 | -10.60 | 705.44 | 0.057 |
| VLDL | -2109.38 | -11659.83 | 7441.06 | 0.661 |
| Total Cholesterol | -239.91 | -573.51 | 93.67 | 0.156 |
| D-Dimer | -0.006 | -0.009 | -0.002 | **0.0012** |
| IFN-Gamma | 1.17 | 1.01 | 1.33 | **<0.0001** |
| IL-6 | -4.85 | -11.59 | 1.87 | 0.155 |
| TNF-a | 0.007 | -0.022 | 0.038 | 0.618 |
| IL-1 | 1.60 | 0.74 | 2.47 | **0.0004** |
| IL-5 | -0.34 | -0.65 | -0.040 | **0.0268** |
| sST2 | -26.01 | -38.15 | -13.87 | **<0.0001** |
| Plasma Potassium | 0.78 | -3.99 | 5.56 | 0.744 |
| Fasting Glucose | 96.00 | -38.00 | 230.01 | 0.157 |
| Lymphocyte Count | -9.70 | -100.43 | 81.02 | 0.831 |
| Microalbumin | 0.91 | -1.15 | 2.97 | 0.382 |

Model 2 was constructed by adjusting for HIV status alone (Table 3). In this model, plasma potassium was a significant positive correlate of IL-17A (β: 10.07, 95% CI: 4.39–15.76, p=0.0006). Additional positive correlates included triglycerides, LDL cholesterol, VLDL, IL-6, TNF-α, IL-1, IL-5, soluble ST2, fasting glucose, and lymphocyte count.

**Table 3.** Multiple linear analysis of factors associated with IL-17A model 2 adjusted for HIV.

| **Parameter** | **B** | **95% L. Limit** | **95% U. Limit** | **P value** |
| --- | --- | --- | --- | --- |
| Triglycerides | 358.64 | 49.67 | 667.61 | **0.0231** |
| LDL Cholesterol | 186.08 | 18.05 | 354.12 | **0.0301** |
| VLDL | 783.73 | 74.99 | 1492.47 | **0.0304** |
| Total Cholesterol | 127.40 | -27.28 | 282.09 | 0.105 |
| D-Dimer | 0.002 | -0.001 | 0.006 | 0.214 |
| IFN-Gamma | 0.043 | -0.005 | 0.092 | 0.081 |
| IL-6 | 21.58 | 15.41 | 27.75 | **<0.0001** |
| TNF-a | 0.03 | 0.01 | 0.06 | **0.0016** |
| IL-1 | 0.021 | 0.004 | 0.038 | **0.0122** |
| IL-5 | 0.38 | 0.15 | 0.62 | **0.0015** |
| sST2 | 15.33 | 5.53 | 25.13 | **0.0023** |
| Plasma Potassium | 10.07 | 4.39 | 15.76 | **0.0006** |
| Fasting Glucose | 28.09 | 15.56 | 240.63 | **0.0259** |
| Lymphocyte Count | 1.09 | 0.30 | 1.89 | **0.007** |
| Microalbumin | 2.45 | -0.51 | 5.42 | 0.104 |

## Discussion

The study findings reveal several independent correlates of IL-17A, highlighting the complex interplay between HIV infection, inflammatory signaling pathways, and metabolic factors. The most robust observation from the primary analysis (Model 1) was the independent association between IL-17A and specific inflammatory mediators, namely IFN-γ and IL-1 (positive correlates), and IL-5, soluble ST2, and D-dimer (negative correlates). Notably, HIV status was not an independent correlate of IL-17A in the model accounting for these inflammatory markers. The association between plasma potassium and IL-17A was model-dependent, emerging as a strong positive correlate only in the analysis adjusted solely for HIV status (Model 2), but not in the comprehensive model containing other inflammatory cytokines.

The strong, independent associations of IFN-γ and IL-1 with IL-17A in Model 1 are biologically plausible. IL-1β is a key cytokine known to drive the differentiation and expansion of Th17 cells, which are the primary producers of IL-17A [12–15]. The concurrent association with IFN-γ, a canonical Th1 cytokine, suggests a possible interplay or shared upstream regulation between Th1 and Th17 pathways in this cohort. The inverse relationships with IL-5 (a Th2 cytokine) and the fibrosis marker soluble ST2 point to a potentially distinct immunoregulatory milieu associated with lower IL-17A levels.

The identification of HIV-positive status as a non-significant predictor in Model 1, but a variable that conditions other associations in Model 2, is an important observation. This finding implies that the influence of HIV infection on IL-17A is largely mediated through or confounded by other inflammatory pathways, consistent with evidence that HIV infection is characterized by persistent immune activation [16]. The novel and context-dependent association between plasma potassium and IL-17A highlights a potentially underrated link that may be masked by broader inflammatory status. Extracellular potassium has been implicated in modulating T-cell function [17] [18], and altered potassium levels might reflect subclinical metabolic or renal disturbances associated with immune activation [19– 21]. This conditional relationship warrants further investigation to clarify its mechanistic basis.

This study possesses several notable strengths that enhance the validity and impact of its findings. First, the analytical approach was rigorous and systematic, progressing from bivariate analyses to multiple linear regression models. This allowed for the identification of independent correlates of IL-17A while controlling for potential confounders. Second, the strategy of building two distinct multivariable models was particularly insightful, as it revealed the context-dependent nature of the association between HIV status and IL-17A, which was masked by other inflammatory cytokines in the full model but emerged as a strong predictor in the clinically oriented model. Furthermore, the study benefited from a comprehensive assessment of a wide array of variables, including detailed sociodemographic, clinical, metabolic, and inflammatory biomarkers, providing a holistic view of potential drivers of IL-17A. The cohort itself is highly relevant to contemporary clinical practice, as the vast majority of participants were on modern INSTI-based antiretroviral regimens, making the findings directly applicable to current populations of people living with HIV.

Several limitations should be acknowledged. The cross-sectional design limits causal inference. The cohort’s demographic composition, predominantly female and largely receiving a specific ART regimen, may restrict the generalizability of these findings. It is also important to note that while other potential confounders such as renal function like eGFR and dietary intake were considered in the initial analysis, they were not statistically significant in the models and were therefore not retained in the final regression tables presented. While this strengthens the case for the identified variables, it does not entirely preclude the possibility of more complex, unmeasured interactions.

## Conclusion

This study demonstrates that in a comprehensive model, IL-17A levels are independently associated with specific inflammatory cytokines (IFN-γ, IL-1, IL-5) and markers (sST2, D-dimer). HIV status was not an independent correlate when these factors were considered. The link between plasma potassium and IL-17A was significant only in a model excluding other inflammatory markers, unveiling a potentially novel and context-dependent pathway in immune-metabolic regulation that merits further longitudinal and mechanistic studies. Understanding these relationships could provide new insights into the chronic inflammation seen in people living with HIV and identify potential targets for immunomodulatory interventions.

## Data Availability

Data is available as supporting information S2

## Availability of data and materials

The raw data underlying the results presented in the study have been uploaded as supporting information.

